# The measurement properties and acceptability of a new parent-infant bonding tool (‘Me and My Baby’) designed for use in a universal healthcare setting in the UK

**DOI:** 10.1101/2021.12.18.21267049

**Authors:** T. Bywater, A. Dunn, C. Endacott, K. Smith, P. A Tiffin, S. Blower, M. Price

## Abstract

NICE guidelines acknowledge the importance of the parent-infant relationship for child development but highlight the need for further research to establish reliable tools for assessment, particularly for parents of children under one year.

This study explores the acceptability and psychometric properties of a co-developed tool, ‘Me and My Baby’ (MaMB).

**Study design:** A cross-sectional design was applied. The MaMB was administered universally (in two sites) with mothers during routine 6–8-week Health Visitor contacts. The sample comprised 467 mothers (434 MaMB completers and 33 ‘non-completers’). Dimensionality of instrument responses were evaluated via exploratory and confirmatory ordinal factor analyses. Item response modelling was conducted via a Rasch calibration to evaluate how the tool conformed to principles of ‘fundamental measurement’. Tool acceptability was evaluated via completion rates and comparing ‘completers’ and ‘non-completers’ demographic differences on age, parity, ethnicity, and English as an additional language. Free-text comments were summarised. Data sharing agreements and data management were compliant with the General Data Protection Regulation, and University of York data management policies.

**Results:** High completion rates suggested the MaMB was acceptable. Psychometric analyses showed the response data to be an excellent fit to a unidimensional confirmatory factor analytic model. All items loaded statistically significantly and substantially (>0.4) on a single underlying factor (latent variable). The item response modelling showed that most MaMB items fitted the Rasch model. Item reliability was high (0.94) yet the test yielded little information on each respondent, as highlighted by the ‘person separation index’ of 0.1 (=>2.0 is required to reliably discriminate between two groups).

**Conclusions and next steps:** MaMB reliably measures a single construct, likely to be infant bonding. However, further validation work is needed, preferably with ‘enriched population samples’ to include higher-need/risk families. The MaMB tool may benefit from reduced response categories (from four to three) and some modest item wording amendments. Following further validation and reliability appraisal the MaMB may ultimately be used with fathers/other primary caregivers and be potentially useful in research, universal health settings as part of a referral pathway, and clinical practice, to identify dyads in need of additional support/interventions.

## 1 Introduction

As mothers are typically primary caregivers, the current study evaluated the MaMB for use by mothers. Maternal bonding can be defined as a mother’s emotional connection and feeling towards her child (Condon, 1993). Bonding is often conflated with attachment. Whilst the constructs are related, they are distinct (Bowlby, 1982; Redshaw and Martin, 2013). Maternal bonding refers to a mother’s (typically self-reported) emotional connection and feelings towards their child. Attachment on the other hand, refers to an infant’s expectations of their caregiver’s responses and the pattern of their own behaviour, e.g., when activated in response to a perceived threat. Attachment typically develops from six months, whereas a mother’s bond to the infant begins to develop during pregnancy. Stronger bonding is theoretically linked to more frequent expression of behaviours such as maternal sensitivity and emotional availability (Feldman et al., 1999), which in turn foster positive interactions within the dyad and promote social and emotional development, including the development of secure attachment in the infant (Ainsworth et al., 1978; Le Bas et al., 2019).

Two systematic reviews (Branjerdporn et al., 2017; Le Bas et al., 2019) indicate that strong maternal bonding in pregnancy is associated with optimal child developmental outcomes. The Le Bas et al. (2019) review also suggested that higher affective postnatal parent-infant bond was predictive of positive child development outcomes. Both reviews suggested the findings should be interpreted with caution due to the relative paucity of studies in this area and highlighted the need for more robust self-report measures of bonding.

There are currently no agreed, standardised, methods for identifying mother/parent-infant dyads who may benefit from additional support around bonding and relationships in England. Although Health Visitors (HVs) work directly with parents some research suggests that they may struggle to consistently identify problems in the parent-infant relationship (Appleton et al., 2013; Elmer et al., 2019; Kristensen et al., 2017; Wilson et al., 2010). Relevant NICE guidelines acknowledge the importance of parent-infant relationship for child development and parent mental health but highlight the need for further research to establish reliable tools for assessment, particularly for parents of children under the age of 1 year (NICE, 2012, 2015).

There is a distinct need for validated, robust measures to be administered universally to identify and support families who may struggle with their parent-infant relationship. Parent-infant relationship is a key focus in the Early Years High Impact Area 2: supporting good parental mental health (PHE, 2020) due to the risks to subsequent child social and emotional development arising from poor parent-infant relationships (Cassidy et al., 2013; Fearon et al., 2010). A reliable, valid, identification tool could allow services to more confidently signpost parents who may benefit to one of the emerging evidence-based interventions (Barlow et al., 2010; Barlow et al., 2016; Facompre et al., 2018; Wright et al., 2015).

A very limited number of brief parent self-report tools exist that assess maternal-infant bonding, are freely available, and have some reliability and validity (Blower et al., 2019; Gridley et al., 2019; Kane, 2017; Wittowski et al., 2020), for example; Maternal Attachment Inventory (MAI; Müller, 1994); Maternal Postnatal Attachment Scale (MPAS) (Condon and Corkindale, 1998); Postpartum Bonding Questionnaire (PBQ) (Brockington et al., 2006); Mother Infant Bonding Scale (MIBS) (Taylor et al., 2005). However, most are not widely used, or have been validated with a small sample (for further discussion see Wittowski et al., 2020; Le Bas et al., 2019). A further two reviews, Blower et al., 2019 and Gridley et al., 2019 were undertaken to explore which measures would be acceptable, reliable, and valid for a large randomised controlled trial of a parenting intervention for parents of infants and toddlers and it was found that choice of measures was very limited (the trial was led by TB, the first author. For the protocol see Bywater et al., 2018).

The 19-item MPAS, which has preliminary evidence of reliability and validity (Kane, 2017; Wittowski et al., 2020) is the most used tool when linking maternal-infant bonding to later child development outcomes (Le Bas et al., 2019). The MPAS was piloted (with the involvement of the first and second authors) with 347 mothers in universal health visiting services (Dunn et al., submitted; Bird et al., submitted) as part of Better Start Bradford - a 10-year National Lottery Community Fund project aimed at improving the socio-emotional development, nutrition and communication skills of children aged 0-3 living in deprived multi-ethnic communities (Dickerson et al., 2016). The pilot concluded that the MPAS could not be recommended for use in health visiting services in Bradford to assess parent-infant relationship due to; little variation in the responses of the 225 who completed the MPAS in English; an unexpected ceiling effect; issues with scoring, parental acceptability and understanding. The E-SEE trial found similar findings, with lack of variation in scores on a sample of 341 (Bywater et al., 2021 (submitted)). Using the learning from the MPAS pilot the study team co-developed a new tool, “Me and My Baby” (MaMB), in an iterative process via workshops and interviews with Health Visitors, Clinical Psychologists, service staff, Managers and parental input, to address the issues highlighted in the MPAS pilot. Prior to a measure being recommended for use in any context, evidence of the measurement properties should be established (Cooper, 2019). Psychometric properties comprise two overarching dimensions - validity and reliability. Validity is defined as the degree to which an instrument measures the construct(s) it purports to measure, and reliability is the degree to which a measure is free from measurement error (de Vet et al., 2015). Acceptable reliability is thus a necessary, though not sufficient, condition for achieving valid scores from an instrument. ‘Reliability’ also relates to the important concept of ‘test information’; that is, the trait level at which the instrument is most capable of discriminating between test takers/respondents. Thus, a test’s ‘information curve’ has important implications for how it is optimally used in practice; for example, when identifying a screening cut-off score.

This study was therefore intended to evaluate the measurement model for the MaMB and acceptability when implemented in routine practice, as a prerequisite to further studies aiming to establish validity of the tool. The main aim was to address previous paucity and quality of available tools to assess parent (mother)-infant relationship, specifically bonding, by developing a measure for use in research as well as universal health settings as part of a referral pathway, and potentially clinical practice, to identify dyads in need of additional support or interventions. The research objectives for this study were:

1. To explore MaMB pilot data to determine the item and test properties in relation to dimensionality and reliability, in terms of both internal consistency and test information; and
2. To identify any necessary revisions to MaMB following the results of our psychometric analysis.

These findings would have implications for which items would be retained in a final version of instrument, and how the scores might be best summarised and used in practice. The work also paves the way for validation studies.

## 2 Materials and Methods

### The tool under investigation

The MaMB questionnaire (for further information see Appendix 2, and the protocol at https://osf.io/q3hmf/) has 11 items presented in a user-friendly format. Responses are indicated using a four-point Likert scale (‘never’, ‘sometimes’, ‘often’, or ‘always’, scored 0-3 with four reversed scored items). The language of items is simple to understand with a reading age of approximately 12, similar to that for popular magazines. A free text box is also included to give mothers the opportunity to record any comments or concerns they have about their relationship with their infant. Lower scores indicate a stronger affective bond.

### Research questions

RQ1: Is the MaMB acceptable to mothers of infants (aged 6-8 weeks) and HVs when administered in a universal healthcare setting?

a. As a proportion of all eligible dyads, how many complete the MaMB?
b. What are the reasons given for non-completion?
c. Are the free text boxes completed by parents and what information is being recorded/reported in them?

RQ2: What are the measurement properties of the MaMB?

a. What is the most plausible dimensionality (factor structure) of the MaMB?
b. Does the scale (or subscales if applicable) of the MaMB demonstrate acceptable levels of internal consistency?
c. According to item response modelling, do the items demonstrate an acceptable fit to the Rasch model, implying that the summed scores from the instrument can be used as a ‘sufficient summary statistic’?
d. What is the relative level of information yielded for respondents by the test (or putative scales), and where might a potential cut-off score be best placed that most accurately differentiates between two groups of test-takers?

### Design

A cross-sectional design was applied.

A briefing was prepared in partnership with Rotherham Doncaster and South Humber NHS Foundation Trust (RDaSH) to support the training of HVs in the use of the tool. The briefing covered the purpose of the tool, how to introduce it to families, how to score it and how to interpret the scores.

The MaMB was implemented universally (in two RDaSH localities) with eligible mothers during the 6–8 week routine HV contact following completion of the core mandated elements of the visit.

HVs asked mothers to complete a paper version of the tool, with support if needed or requested. During tool completion HVs were expected to use their professional skills to discuss with parents their relationship with their infant. If HVs were unable to complete the tool (e.g., due to time constraints) they would record the reason(s) for non-completion.

HVs inputted the responses electronically into the case management software (SystmOne) co-developed template to include; if tool administration was attempted, and if not why, and if tool administration had been abandoned prior to completion. The template also captured responses to all 11 items, and the free text responses to the open question on the back page of the paper tool, and HVs comments on the interaction. Key demographic variables were also recorded to adequately describe the sample’s characteristics and to support subgroup analyses.

The research team received anonymised (numerical and free text) data extracted from SystmOne, and a small number of key demographic characteristics such as age, ethnicity, and parity.

### Study setting

Two RDaSH sites in Northern England implemented the MaMB at the 6-8 week universally mandated HV contact.

### Inclusion/exclusion criteria

All mothers of a child aged 6-8 weeks living in the sites were eligible for the study.

If a parent had opted out of NHS digital they may have completed the MaMB but were not included in the study (in England, NHS patients can choose to opt out of their confidential patient information being used for research and planning).

### Consent

This study received ethical approval on 21st August 2020 by South Central - Berkshire B Research Ethics Committee, UK, Ref: 20/SC/0266, Integrated Research Application System (IRAS) 201, project ID: 273708.

Parents were given a MaMB Participant Information Sheet (V2.0 17^th^ August 2020; See Appendix 1) at a visit prior to the 6-8-week check to give them time to read and understand why they will be asked to complete the MaMB.

Written consent from mothers completing the MaMB, and for the non-identifiable fully anonymized, data to be shared with the research team, was not required. This was because:

1. The research team only accessed anonymised data. Data were restricted to the minimum needed to describe the sample and to conduct the proposed analyses of measurement properties and acceptability. Free text boxes, where completed, and were screened by an authorised RDaSH employee to remove any identifiable information prior to data sharing.
2. There was no risk of harm to participants from completing the MaMB. The tool was one of several used by HVs to conduct a broad needs assessment, as is standard at the 6–8-week contact. The MaMB supplemented existing tools and was implemented in addition to standard care. HVs are trained and well equipped to support mothers who may be struggling to bond with their baby.
3. It was deemed essential that the MaMB sample was representative of mothers of young infants in the research site so that the study findings are generalisable. Introducing an informed consent process would likely have led to selection bias, arising from parent and practitioner characteristics and attitudes.
4. There is a clear value and benefit from doing the research, i.e., a need for a short, easy-to-administer, valid and reliable measure to support practitioners to identify families experiencing difficulties in their parent-infant relationship. The MaMB has been co-developed by academics, psychologists and HVs with parental input to address this gap, it is vital that this measure is tested before it can be recommended for use more widely.

### Sample size

The average number of live births per year in the year prior to the study was 3460 in Site 1 (Doncaster) and 3000 in Site 2 (North Lincolnshire), which would yield approximately 538 births per month. Assuming a conservative 50% completion rate (allowing for potential implementation/uptake barriers such as time constraints, parent refusal or practitioner non-compliance, time lag in implementation and data entry) we anticipated 269 MaMBs would be completed per month. To construct a sample large enough to support the analysis of psychometric properties we proposed a sample of 673 over a ten-week period. Based on a 50% completion rate, the overall sample would include a further 673 non-completers to explore acceptability (total n=1346). Please note this sample size was calculated pre-COVID-19.

### Psychometric analyses RQ1

To assess acceptability of the tool reported the proportion of participants who were recorded as being offered the tool but either refused, or failed to complete, it. Where data were available descriptive analysis of the reasons for refusal was to be produced.

Key demographic characteristics (age, parity, ethnicity, English as an additional language) of completers and non-completers were to be presented in contingency tables as either frequency counts or means for descriptive purposes.

A frequency count was intended to determine the proportion of completers who used the free-text box to expand on their answers. Free-text comments were to be summarised in a brief narrative.

### RQ2

#### Dimensionality and internal consistency reliability

The sample was originally intended to be randomised into exploratory and confirmatory (‘validation’) datasets, if the data obtained were sufficient to support this approach. Initially dimensionality was planned to be explored in the former data subset using parallel analysis (see below for details) (Horn, 1965). Once this had been established, it would be followed by an exploratory factor analysis (EFA) of exploratory portion of the response data. The potential factor structures elicited would then be tested using confirmatory factor analyses (CFA) on the confirmatory (validation) dataset. Internal reliability consistency of the postulated subscales would then be examined. The findings of these analyses were intended to indicate whether it is appropriate to summarise bonding via several subscales or simply by a single total overall score for the MaMB.

The parallel analysis would be performed using unweighted least squares (ULS) as the estimation method (Horn, 1965; Lorenzo-Seva and Ferrando, 2006). In a parallel analysis the maximum plausible number of factors to be retained is indicated at the point where the eigenvalues of the randomly generated data exceed those of the actual data. A series of EFAs was expected to be then performed to aid interpretation of any factors underlying the response patterns observed. Oblique (geomin) rotations were to be used in the factor analyses, assuming that, as in almost all psychological measures, underlying latent traits would be correlated with each other to some extent. The EFAs will be repeated, again using a geomin rotation, to derive standard errors (and thus standardised Z scores) for the factor loadings to evaluate their relative statistical significance (Asparouhov and Muthén, 2009). All EFAs and CFAs were to be conducted in Mplus version 6.1 employing robust weighted least squares (WLSMV) as the estimation method, or ‘full information maximum likelihood’, as appropriate.

Internal reliability consistency for the putative subscales based on the CFA structure was to be evaluated using Cronbach’s alpha and McDonald’s omega. Cronbach’s alpha may be a poor index of internal reliability where tau-equivalence (equality of factor loadings across items in a scale) does not hold (Raykov, 1997). In this respect McDonald’s omega is reported to represent a more accurate estimate of the extent to which items in a scale measure a unidimensional underlying construct.

#### Item response modelling

Item response modelling and theory (IRT) is based on the modified factor analysis of binary and categorical data. Within the family of IRT models Rasch analysis was originally developed for the exploration of dichotomous responses to test items (Rasch, 1992), though was subsequently extended to accommodate polytomous data. Rasch analysis can be used to create interval metrics of both item difficulty and respondent ability from ordinal (ordered categorical) or binary (dichotomous) response data. The Rasch model assumes that all items are identical in terms of their ability to discriminate between respondents according to ability/trait (i.e., equality of item factor loadings in classical factor analytic terms). Nevertheless, Rasch software can provide simulated estimates of other parameters aside from difficulty and ability such as the degree of discrimination an item provides in determining the level of the underlying trait in a respondent. In a Rasch analysis reliability can be appraised in several ways. Specifically, the person reliability coefficient relates to the replicability of the ranking of abilities while the person separation index represents the signal to noise ratio and estimates the ability of a test to reliably differentiate different levels of ability within a cohort (Wright and Masters, 1982).

Power issues in Rasch analysis are a matter for debate with some authors suggesting that around 200 respondents are required to accurately estimate item difficulty whilst others suggest as few as 30 participants may be required in well-targeted tests (i.e. those where difficulty is well matched to ability) (Baur and Lukes, 2009; Goldman and Raju, 1986; Linacre, 1994).Thus, this study should be adequately powered to estimate item properties from both Rasch analysis as well as the factor analyses, the latter of which could be considered re-parameterized two parameter logistic regression IRT models. Thus, the fit of items to the Rasch model was to be assessed and any potential sources of misfit diagnosed. This will be important in deciding whether it is appropriate to summarise the scores on the scale/s as summed totals. Moreover, the Rasch calibration was intended to allow the evaluation of test information, which would indicate to what extent the test is able to differentiate test-takers across the putative trait levels under evaluation (assumed to be ‘perceived bonding with baby’).

### Data handling and sharing

Fully anonymised data was exported from SystmOne and shared with the study team via the University of York secure drop off service, which securely encrypts data. Data management is compliant with the General Data Protection Regulation (GDPR) and University of York data management policies. The custodian of data, Professor Tracey Bywater (Chief Investigator), is the contact point for any data management queries.

## 3. Results

The pilot ran 10^th^ Sept 2020 to 1^st^ Dec 2020, and the MaMB was administered either face to face or over the telephone depending on COVID-19 restrictions at the time of administration.

During the pilot:

- Women due 6-8-week maternal contact = 1,031
- Women had a 6-8-week maternal contact = 928
- Women with completed MaMB = 434

The 434/928 equates to a 47% response rate, close to the 50% we predicted.

The sample size was lower than originally proposed of N=673, and of the 494 women who did not complete the MaMB we only have data for 33 women rather than the proposed 673.

Results will be presented in order of the research questions.

**RQ1:** Is the MaMB acceptable to mothers of infants (aged 6-8 weeks) and HVs when administered in a universal healthcare setting?

a. As a proportion of all eligible dyads, how many complete the MaMB?
b. What are the reasons given for non-completion?
c. Are the free text boxes completed by parents and what information is being recorded/reported in them?

Table 1 shows the characteristics of the sample who completed the MaMB, and the sample appears to represent the local population regarding ethnicity and language. Although the numbers are small and we cannot draw conclusions from them, the 33 non-completers appeared to differ on ethnicity and language, which may be a reason for not completing the MaMB, e.g., 24% were white ‘other’ in the non-completers, compared to 10% in the completers. Likewise, 38% of non-completers needed an interpreter compared to 14% from the completers. Although 461 cover sheets for non-completers were missing, there was minimal missing data at item-level for those that were returned.

**Table 1.** Characteristics of completers (N=434) and non-completers (N=33)

|  |  | **Completers (N=434)** |  | **Non-Completers (N=33)** |  |
| --- | --- | --- | --- | --- | --- |
|  |  | Count | Percent | Count | Percent |
| Site |  |  |  |  |  |
|  | Doncaster (Site 1) | 256 | 59% | 21 | 64% |
|  | North Lincolnshire (Site 2) | 178 | 41% | 12 | 36% |
| Mother age (in years) |  |  |  |  |  |
|  | Mean (SD) | 28.45 (5.76) | / | 29.25 (5.17) | / |
|  | Min | 16 | / | 21 | / |
|  | Max | 43 | / | 43 | / |
| Child age (in weeks) |  |  |  |  |  |
|  | Mean (SD) | 6.69 (1.69) | / | 8 | / |
|  | Min | 4 | / | 6 | / |
|  | Max | 25 | / | 31 | / |
| Ethnicity |  |  |  |  |  |
|  | White British | 359 | 83% | 16 | 49% |
|  | White Other | 43 | 10% | 8 | 24% |
|  | Asian/Asian British | 13 | 3% | 0 | 0 |
|  | Black African | 5 | 1% | 3 | 9% |
|  | /Caribbean/Black British |  |  |  |  |
|  | Mixed/Multi-ethnic | 2 | 0.5% | 1 | 3% |
|  | Other | 9 | 2% | 1 | 3% |
|  | Missing | 3 | 0.5% | 4 | 12% |
| Mother's first language is English |  |  |  |  |  |
|  | Yes | 348 | 80% | 15 | 46% |
|  | No | 59 | 14% | 13 | 39% |
|  | Missing | 27 | 6% | 5 | 15% |
| Interpreter needed (for non-first lang English) |  |  |  |  |  |
|  | Yes | 8 | 14% | 5 | 38% |
|  | No | 50 | 85% | 7 | 54% |
|  | Missing | 1 | 1% | 1 | 8% |
| First child |  |  |  |  |  |
|  | Yes | 195 | 45% | 9 | 27% |
|  | No | 235 | 54% | 20 | 61% |
|  | Missing | 4 | 1% | 4 | 12% |

N.B. Table 1 includes a descriptive summary of available data from the 33 women who did not complete a MaMB but their health visitor completed a cover sheet

From the 434 respondents who completed a MaMB 50 had one or more missing items. The scores for the 384 who fully completed MaMBs suggest that the sample had positive relationships with their baby, mean = 1.2 (SD 1.6) from a possible 33 (the lower the score indicating the more positive the perception of the mother-baby relationship), and a range of 0-15. Twenty-nine respondents (parents and HVs) completed the free-text box with some mothers saying they felt guilty that they could not give more time to their baby or felt less than positively to toward their child at times, e.g;

> *“I feel guilty for having less positive feelings especially when he is screaming” “I feel I need time by myself sometimes, but feel guilty that I feel like that as a mum”*
>
> Four mothers mentioned that they had not been separated from their baby yet, so items 8 and 10 were not applicable.
>
> **RQ2:** *What are the measurement properties of the MaMB?*

From 471 mothers 33 had no MaMB questionnaire data, leaving 438 participants with some response data. The original plan was to divide up the data, randomly, into a training and validation set (see Methods). However, due to lack of variance in some of the item responses this was not possible. That is, dividing the dataset into two portions created items where little or no variation in responses were observed in some cases, rendering estimation of factor models impossible. Therefore, the entire dataset was explored in relation to its dimensionality.

### Dimensionality

Firstly, a parallel analysis was conducted using the software FACTOR. This generates pseudorandom data, with the same dimensions as the real data. This process was adapted for use with the ordinal response data using polychoric matrices. Missing data values were handled using hot deck multiple imputation (Lorenzo-Seva and Van Ginkel, 2016). The results of the parallel analysis are shown in Table 2. These clearly indicate that there is a maximum of one factor (latent variable) underlying the response structure. This is evidenced clearly by the fact that the first latent variable explains around 50% of the variance in the indicators (item responses). However, a second postulated latent variable explains less variance than that found in a second latent variable for the pseudorandom data. The reliability, as indexed by Cronbach’s alpha was 0.64, though the alpha-KR20 (adapted for binary or ordinal data) was 0.88 (as calculated by Winsteps-see below).

**Table 2.**
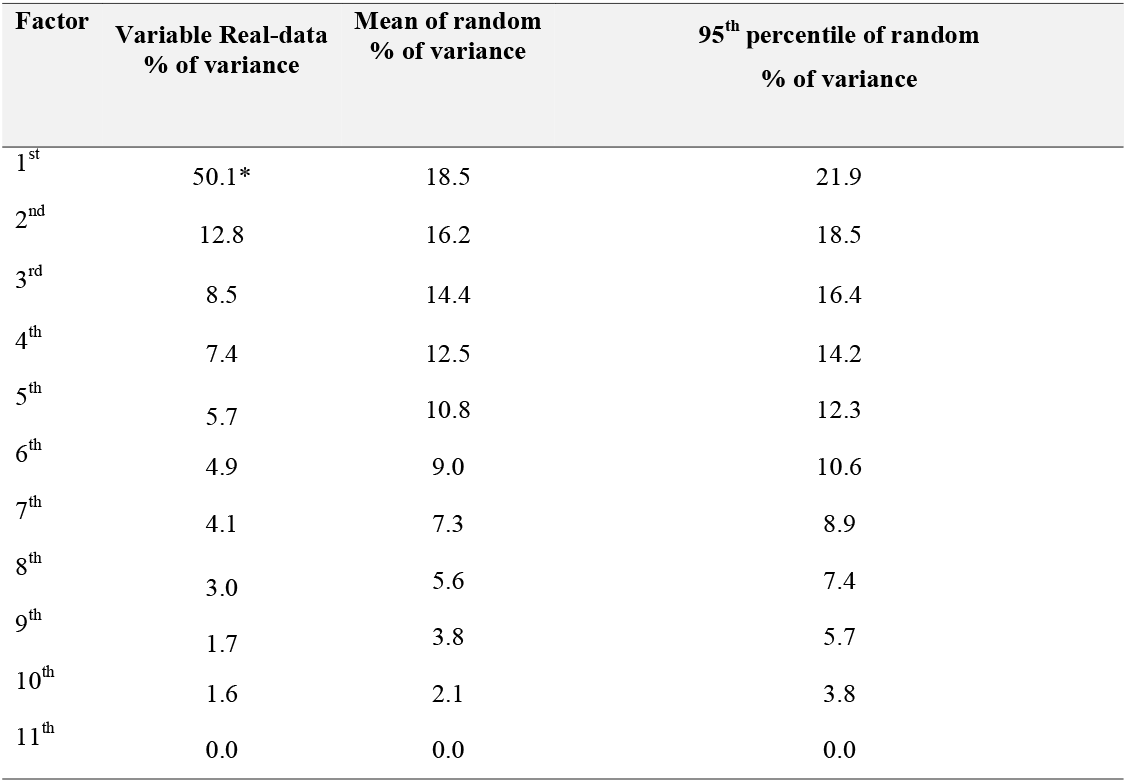
Results from a parallel analysis, adapted for ordinal data. *Note only the percentage of variance explained by the first factor exceeds that observed for the random data.

This unidimensional structure was confirmed by examining the fit to a single factor confirmatory factor analytic model within the Mplus v8.4 software environment. This confirmatory factor analysis (CFA) was adapted for the ordinal nature of the response data, using robust weighted least squares as the estimation method (WLSMV). There were technical difficulties estimating a one factor model due to the low variance in items 4 and 5 and their collinearity with responses to items 10 and 11 respectively (that is, responses to the latter items were almost wholly associated with response to the former). Specifically, the correlation between item 4 (‘difficult’) and item 10 (‘apart’) was 0.987. That between item 5 (‘need’) and item 11 (‘play’) was also 0.987. Consequently, items 4 and 5 (which exhibited the lowest variance of the pairs were dropped from the CFA. When the CFA was repeated with the remaining nine items the one factor model showed an excellent fit to the data; the CFI and TLI fit indices were both 0.99 (≥0.90 usually is taken as acceptable fit, whilst values over 0.95 indicate good fit). The standardised factor loadings are shown in Table 3.

**Table 3.**
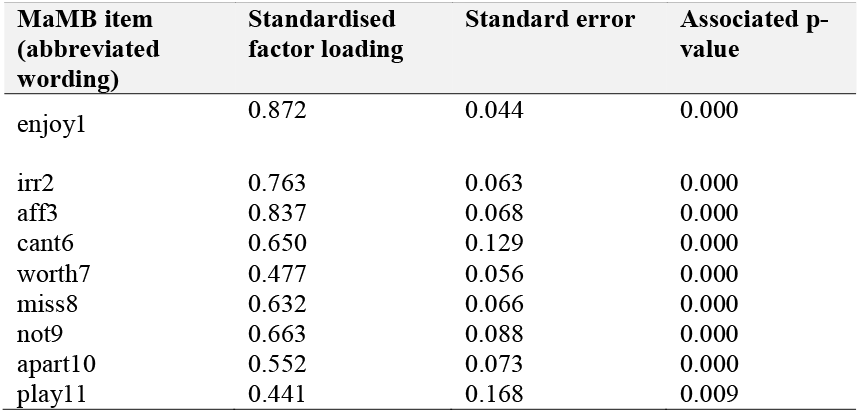
Standardised factor loadings of the MAMB items estimated using a one factor confirmatory [ordinal] factor analytic (CFA) model.

| MaMB item<br>(abbreviated<br>wording) | Standardised<br>factor loading | Standard error | Associated p-<br>value |
| --- | --- | --- | --- |
| enjoy1 | 0.872 | 0.044 | 0.000 |
| irr2 | 0.763 | 0.063 | 0.000 |
| aff3 | 0.837 | 0.068 | 0.000 |
| cant6 | 0.650 | 0.129 | 0.000 |
| worth7 | 0.477 | 0.056 | 0.000 |
| miss8 | 0.632 | 0.066 | 0.000 |
| not9 | 0.663 | 0.088 | 0.000 |
| apart10 | 0.552 | 0.073 | 0.000 |
| play11 | 0.441 | 0.168 | 0.009 |

The factor loadings demonstrate a substantial (>0.4), positive and significant (p<0.01) magnitude of loadings for all nine MaMB items included. Negative items were reverse coded so that the latent variable and the item factor loadings were interpretable. Having established the unidimensional structure of the data it appeared appropriate to progress to a Rasch calibration of the MaMB items.

### Rasch analysis

The Rasch calibration results yielded much useful diagnostic information on the MaMB questionnaire. As highlighted earlier the scale reliability itself was moderate to high. Indeed, the item reliability estimated by the Rasch calibration was .94. However, the person separation index (which include ‘extreme’ and ‘non-extreme’ persons) was only .10. The person separation index reflects the number of groups that can be plausibly differentiated by the scale with acceptable precision. It represents a signal to noise ratio in the scale. This should normally exceed 2.0. Thus, the MaMB scale had virtually no ability to differentiate respondents. This was no doubt a reflection on the lack of observed variance in responses in the study sample. Nevertheless, in terms of scale development and future research it is useful to explore the item ‘difficulties’ (or ‘endorsibility’ in this case), as well as the fit statistics. These are shown below in Table 3. The z standardised fit, along with the difficulty/endorsibility and standard error (reflected in the diameter of each bubble) are also shown in the ‘bubble plots’ in Figures 1 and 2. In the Rasch context ‘fit’ in this sense refers to which the item responses follow a Guttman sequence (Rasch, 1960). That is, as the ability or trait increases the respondent or test-taker tends to be observed to give a higher scoring category of response, allowing for the play of chance. E.g., 0010101112221221222223323333. Items where responses are too predictable (e.g., deterministic, 00111222333) ‘overfit’ the model. Those that are more erratic (e.g., 021231313) are described as ‘underfitting’. The former tends to indicate redundant items, the latter, erratic items that can distort or degrade the measurement properties of the scale. ‘Infit’ refers to fit where an item ‘difficulty’ is well matched to the level of trait or ability in a test taker. That is, for example, for a right/wrong maths question the person who is well matched would have a 50:50 chance of either a correct or incorrect answer. In this case ‘well targeted’ items would tend to show a reasonable spread of responses for a set of test takers with trait levels that are matched to the item endorsibility. Conversely, ‘outfit’ refers to fit (conformity to the Rasch model) where item difficulty is not well matched to the test taker’s trait or ability level. These distinctions between infit and outfit tend to be more pertinent to knowledge tests, than trait assessments, however. As can be seen from Table 4 and Figures 1 and 2, overall, the MaMB items tend to conform reasonably well to the Rasch model. However, there are four key issues.

**Table 4.** Item ‘endorsibility’ (‘measure’) of the MaMB scale, along with the Rasch fit statistics. These include both ‘infit’ and ‘outfit’ statistics as both the mean squared error and standardised (z) <u>fit.</u>

| Item | Item difficulty/<br>‘Endorsibility’ | Infit<br>(mean-squared) | Infit<br>(standardised ) | Infit (mean-squared) | Outfit<br>(standardised ) |
| --- | --- | --- | --- | --- | --- |
| enjoy1 | -0.7 | 0.7557 | -1.9392 | 0.5332 | -2.3195 |
| irr2 | 0.73 | 0.8987 | -0.7791 | 0.6421 | -1.5994 |
| aff3 | -0.37 | 0.8241 | -0.5392 | 0.539 | -0.7295 |
| diff4 | -0.8 | 0.9757 | 0.001 | 1.3484 | 0.7613 |
| need5 | 2.51 | 1.1986 | 2.0112 | 1.3104 | 2.6813 |
| cant6 | 0.09 | 1.0214 | 0.241 | 0.9482 | -0.4291 |
| worth7 | -0.32 | 1.0671 | 0.3511 | 1.2485 | 0.6412 |
| miss8 | -0.15 | 1.0677 | 0.4411 | 1.0662 | 0.3311 |
| not9 | -0.17 | 1.1529 | 0.5212 | 2.1197 | 1.3821 |
| apart10 | -0.47 | 0.9778 | 0.071 | 0.9267 | 0.0709 |
| play11 | -0.34 | 0.8633 | -0.4391 | 1.1688 | 0.4812 |

**Figure 1.**
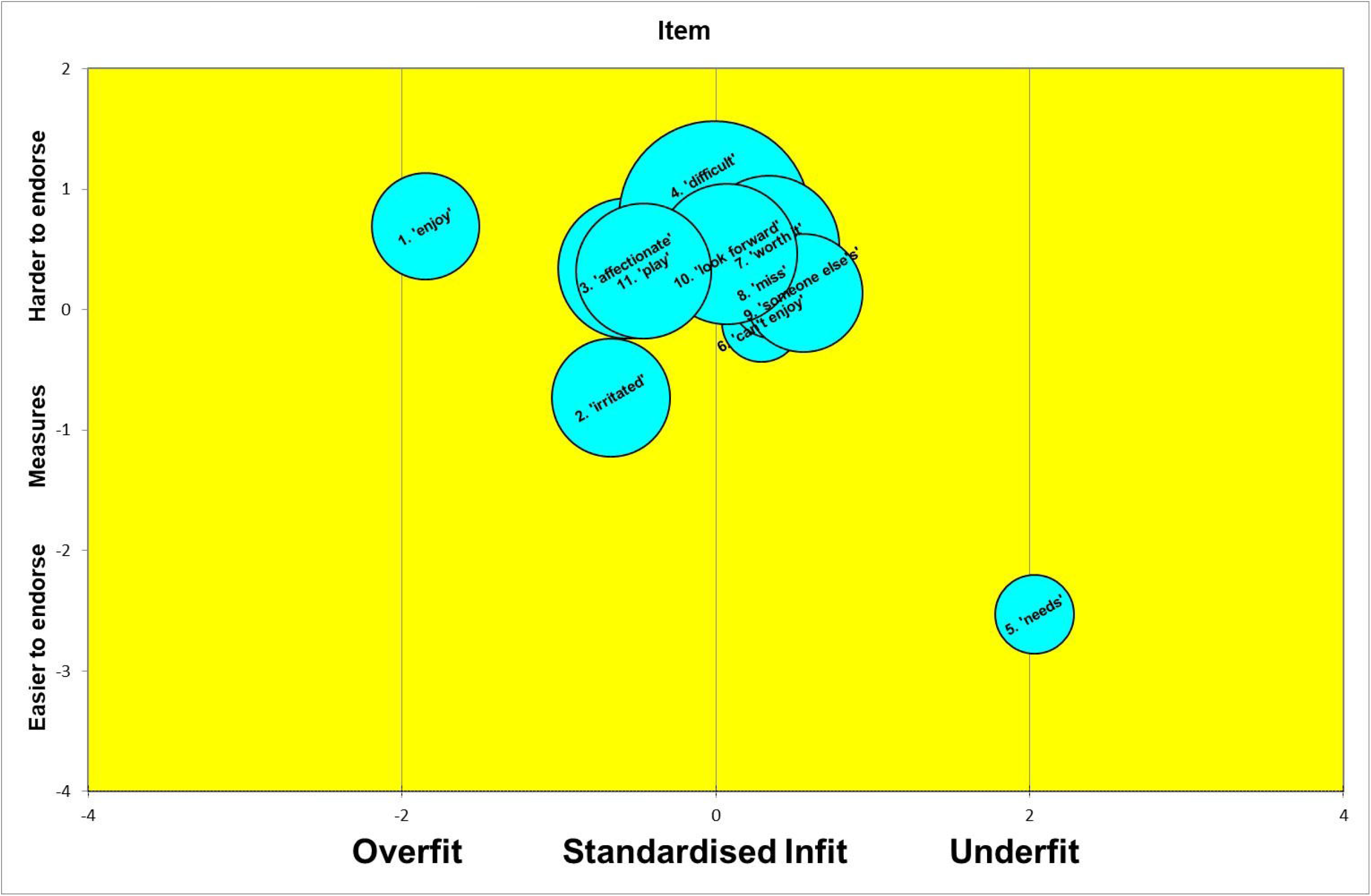
Bubbleplot

**Figure 2.**
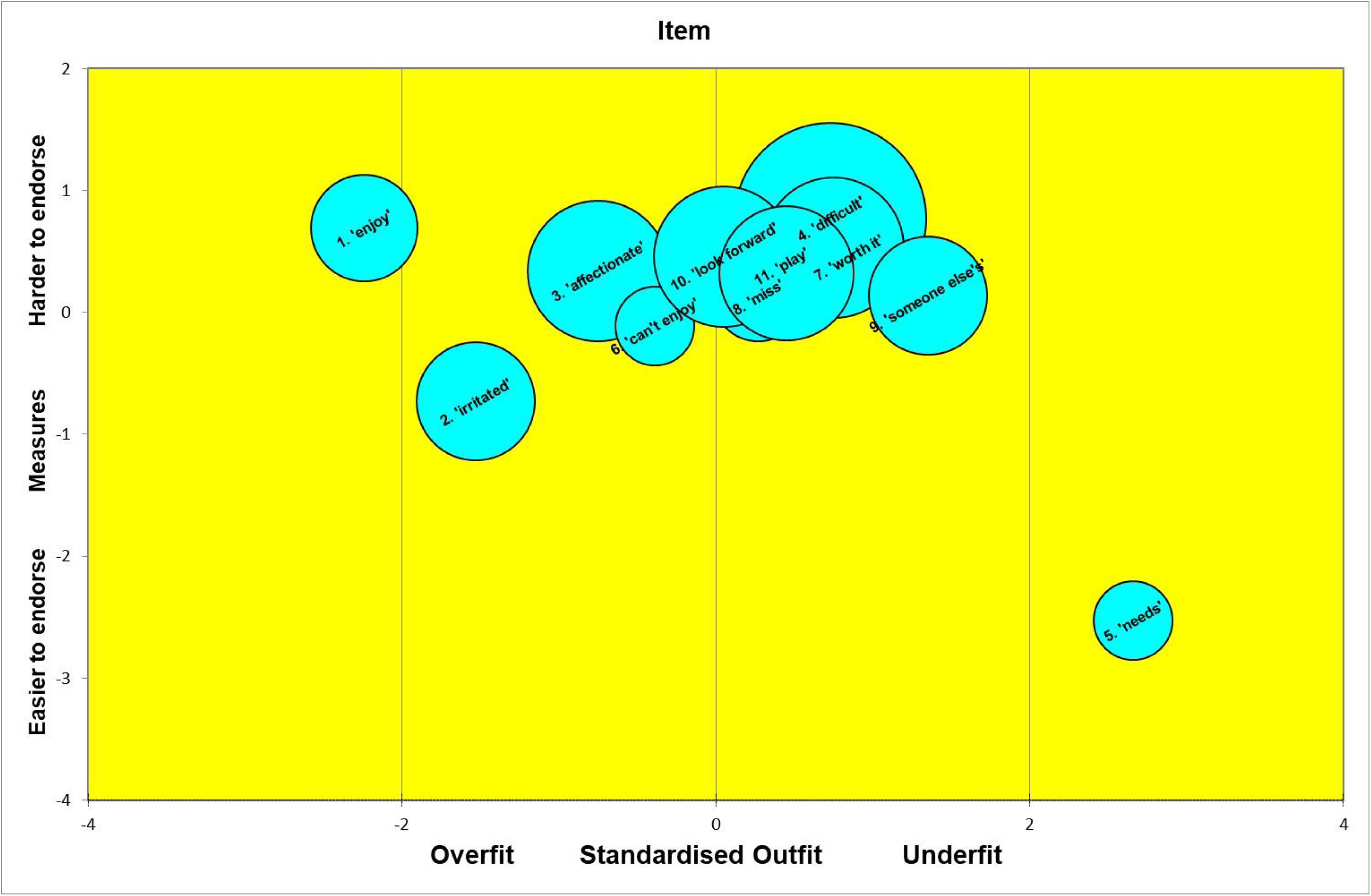
Bubbleplot

1. The items seem very easy (or in the case of negatively worded items-very hard) to endorse. This can be seen by the ‘measure’ estimates that tend to be around or below the zero point-a standardised trait (estimate) derived from the item responses.
2. A couple of items tend to overfit the model: ‘enjoy’ (item 1) and ‘irritated’ (item 2)). These tend to be somewhat overly predictable from the responses to the other items. However, this observation should be viewed cautiously as only the z standardised fit showed misfit, and this can be sensitive to relatively large numbers of observations (e.g. >300).
3. One item (‘not mine’ – item 9) tends to show poor outfit. This suggests some erratic ratings, by those respondents whose estimated trait level was some distance from the item ‘measure’ (endorsibility’) of −0.17.
4. One item showed poor infit and outfit, at least on the ‘z’ fit statistics (‘needs’). This suggests this item may have been relatively erratically answered. It may have been different respondents read or interpreted the item differently from each other. For example, some may have interpreted it in terms of basic needs, whilst others, more in terms of emotional needs. It may be useful to explore whether this item showed any item bias or differential item functioning in relation to demographic factors.

### Item category probabilities

It was apparent that most of the items were not operating as four-point Likert scales. That is, in many items not all four categories of response were observed in this sample of respondents. Moreover, some intermediate categories of response were rarely observed. In effect this means that even if a respondent is higher on a trait level a lower category of response may still be observed. This is sometimes referred to as ‘Rasch-Andrich threshold suppression’. This effect is nicely illustrated below, by the item category probability curves for item 11. Although some respondents selected a response with a score of ‘2’ had higher trait levels than those who scored ‘1’ (‘0’ was not observed), in practice they were more likely to be seen to choose a ‘1’ category, as so few chose the ‘2’. These findings suggest, at least for the kind of general population sample used in this study, the use of four Likert scale points may be too many; that is, they may not lead to more information on a test-taker and also introduce some risk of extreme responses style (ERS) bias. *Figure S1*, in the supplementary material, refers to probability of observing a respondent choosing a particular response category according to their overall trait level (‘baby bonding’). Note that curves do not always correspond to the ordered responses (0->1->2→3).

### Test information

As would be expected for a test mainly composed of easily endorsed items, most of the area under the test information curve was for test takers whose traits were defined as slightly below the average. That is, those who were likely to give midrange responses to easily endorsed items. This can be seen by the fact the peak of the test information curve is just below the zero on the x-axis. This suggests the item calibration is not ideal to pick out mothers who may be struggling to bond with their babies (i.e., those who are likely to be observed with a lower total score on the MaMB scale). The test information curve is depicted in *Figure S2* in the provided Supplementary Material.

## Discussion

There is a paucity of high-quality tools to assess parent-infant relationships. The MaMB was co-developed to address this gap and act as a tool to measure bonding for use in research and universal health settings.

The results suggest that it is feasible for HVs to administer the MaMB with mothers in universal services. HVs successfully completed the MaMB with approximately 50% of the universal population at the 6-8-week visit in the context of highly pressured services due to the Covid-19 global pandemic. Given low rates of missing data the MaMB appears to be acceptable to parents.

The psychometric analyses suggest the MaMB tool responses, in this sample of test takers, were unidimensional. The MaMB showed relatively high levels of internal reliability consistency and the items generally fitted the Rasch model. However, the high reliability may be partly an artefact of the lack of variation in responses observed – almost all respondents gave high-scoring categories on the items. The items did not generally behave as four-point response format questions, as it was common for some response categories to go unobserved. Consequently, test information was relatively low and was much less than may be required to identify at least two separate groups of respondents, e.g., if the MaMB were to be used as a screening tool.

For the 29 parents that completed the free text it appeared a useful part of the MaMB to expand on item completion with an opportunity to voice feelings or concerns. Responses suggest parents were engaging in a meaningful discussion about bonding with their health visitor. This suggests the MaMB could be considered a potential catalyst in opening discussions about sensitive aspects of parenting such as experiencing guilt for wanting some ‘alone’ time, or for feeling less positive when their baby is screaming. Such open conversations suggest that the tool could fit well within a pathway for accessing specialist services, such as infant mental health services.

### Strengths

The MaMB was co-developed over a series of workshops and interviews, using an iterative process with HVs, Clinical Psychologists, service staff and managers from different localities, and included parental input. It was piloted within routine HV contacts and, although the pilot was delivered during the COVID-19 pandemic with many visits taking place remotely, or with restrictions, completed MaMBs were obtained from 50% of the potential population. The pilot study was classed as research as opposed to service design and had ethical approval as such. Previously psychometric analyses focused on exploratory and confirmatory factor analysis; however this study also included IRT, which affords additional rigour and confidence in the results.

### Limitations

Some HV teams would have conducted some core 6-8-week contacts over the telephone rather than in the family home due to COVID-19. However, we do not have data to report how many. This may have led to lower completion rate of the MaMB.

A much smaller than anticipated comparison group of non-completers was achieved. This was because HVs appeared not to complete, or partially complete, a cover sheet with demographic information if a mother did not wish to complete the MaMB. The pilot was conducted during the COVID-19 pandemic, during which time HVs were under enormous pressure to continue delivering statutory support to families despite adverse circumstances which likely contributed to the non-completion of cover sheets.

### Deviations from the registered protocol

Due to the limited information on non-completers we were unable to conduct planned statistical analyses of the characteristics of completers compared to non-completers. The amount of data contained within the free-text responses of completed MaMBs also prevented a planned thematic analysis of these data, though it was sufficient to provide useful information in a descriptive summary.

### Future research

The findings of this study suggest that the MaMB is a promising tool to assess parent-infant relationships. Future research directions fall across three domains (1) understanding practitioner experiences, (2) expanding sample of users, and (3) refining approach to measurement.

#### Understanding Practitioner Experiences

Practitioners such as health visitors are a key component of using a measure of parent-infant relationships. A better understanding of their experience supporting mothers to complete the MaMB tool would help to further refine the tool. Obtaining ethical approval to ask HVs from the current study their views on completing the MaMB would be a priority for future research.

#### Expanding Sample of Users

This study found that most participants responded similarly to items on the MaMB. Further piloting of the tool with an expanded sample of users would help to understand the reason for this limited range of responses. For example, use with mothers experiencing mental health difficulties in the perinatal period would be particularly valuable. We might hypothesise that those within the clinical range of depression measures may respond differently when asked about their bond with their baby. This is highly likely to result in observing more variance in the items. It may also be able to show whether the tool is able to discriminate, with any precision, between at least two different groups of respondents. Note, in theory, a Rasch model is based on a sample free distribution (that is the estimates should be the same irrespective of the sample of test takers used for the calibration). However, in practice, precise estimates of item fit and difficulty may not be achieved, even with large samples, if some categories of response are rarely or never observed.

It was appropriate for this first pilot to target mothers, who are typically primary caregivers. However, we know that there is increasing variability in those who take on the primary caregiver role across society. Piloting the MaMB tool with a diverse range of caregivers would enable exploration of differences and similarities across responses for wider parent-infant relationships. It would also support use of the tool in practice, where fathers, same sex parents, or other kinship carers may be caring for a baby.

#### Refining Approach to Measurement

To enable the tool to have a greater degree of variation across responses, future research could test the MaMB tool with amended items (as highlighted in the results) to make them more subtle. This could be helpful in picking up difficulties and bonding and attachment in parents or caregivers. Moreover, future research could evaluate the tool as a three-point Likert scale, as opposed to the four-point scale used in the current study. This could help to increase variation across items.

## Conclusions

HVs successfully administered the MaMB in universal services and the MaMB appears to be acceptable to parents. The MaMB demonstrated good internal consistency and may support HV signposting decisions for additional support, however, as the more robust analysis shows, if the MaMB was to be used as a screening tool, with a cut-off, or ranges of ‘concern’ then additional work is needed, which will need to include more families with risk factors such as depression in an enriched sample.

Regarding our objectives, we consider the MaMB to be feasible for use in routine practice with some amendments, and future piloting of such amendments.

## Data Availability

The custodian of data, Professor Tracey Bywater (Chief Investigator), is the contact point for any data management queries

## ASSOCIATED PROTOCOL

Version 2, 7^th^ December 2020 – to access the protocol for further information please visit: https://osf.io/q3hmf/ Bywater, T., Blower, S. L., Dunn, A., Endacott, C., Smith, K., & Tiffin, P. (2020, December 7). Me and My Baby Protocol: The measurement properties and acceptability of a new mother-infant bonding tool designed for use in a universal healthcare setting in the UK. Retrieved from osf.io/6br2e

## RESEARCH REFERENCE NUMBERS

- IRAS Number: 273708
- This study received ethical approval on 21st August 2020 by South Central - Berkshire B Research Ethics Committee, UK, Ref: 20/SC/0266, Integrated Research Application System (IRAS) 201, project ID: 273708.Funder References : HEIF - H0026802, ARC-YH: NIHR200166, The National Lottery Community Fund (previously the Big Lottery Fund) - 0094849, National Institute for Health Research (NIHR) Public Health Research (PHR) (ref 13/93/10).
- OFS Study Registration Number: osf.io/6br2e

## AUTHOR CONTRIBUTIONS

TB secured funding, TB and AD conceived the study, TB, AD, CE, KS, PAT, SB designed various aspects of the study. TB and SB provided supervision of the study from the academic perspective (UoY) and KS from the practitioner perspective (RDaSH). PAT conducted the statistical analysis and provided psychometric expertise, and KS and MP participated in co-developing the tool and provided clinical and practitioner expertise. AD and CE were research fellows on this study and coordinated and conducted various activities.

TB wrote the initial draft, and all authors have contributed and commented on subsequent drafts of this paper. TB, corresponding author, will act as guarantor and affirms that the manuscript is an honest, accurate, transparent, and full account. All listed authors meet authorship criteria and no others meeting the criteria have been omitted.

## ACKNOWLEDGEMENTS

This study could not be conducted without the support of RDaSH, the health visiting teams from Doncaster and North Lincolnshire, and of course the families - we extend our gratitude to these key stakeholders, and to other professionals and parents across Yorkshire and Lincolnshire who participated in the co-development of the MaMB. We would also like to thank the funders (see funding section for information), and the ARC-YH management group for their oversight. The views expressed in this publication are those of the authors and not necessarily those of the NHS, the NIHR or the Department of Health and Social Care. This study also received funding from the National Lottery Community Fund (previously the Big Lottery Fund) as part of the ‘A Better Start’ programme. The National Lottery Community Fund have not had any involvement in the design or writing of the paper.

## SUPPLEMENTARY MATERIAL

Supplementary material is provided for this study.

## DISCLAIMER

The views expressed in this publication are those of the authors and not necessarily those of the NHS, the NIHR or the Department of Health and Social Care. This study also received funding from the National Lottery Community Fund (previously the Big Lottery Fund) as part of the ‘A Better Start’ programme. The National Lottery Community Fund have not had any involvement in the design or writing of the paper.

## Appendix 1: The MaMB Participant Information Sheet (To include logos)

### Information on the ‘Me and My Baby’ questionnaire

#### What will happen at your next Health Visitor visit?

When your baby is between 6 and 8 weeks old your Health Visitor will talk to you about how you and your baby are getting along. At this visit your Health Visitor will ask if you have any questions about the information in this leaflet, and if you’re willing to complete some questions about your relationship with your baby.

#### Why are you asking about my relationship with my baby?

The Me and My Baby questionnaire is short and has been developed by Health Visitors and researchers, with other NHS staff, and with input from parents.

Not everyone finds it easy to get on with their new baby. Some mums, even if they have other children, sometimes feel they don’t understand their new baby, or that their baby is being difficult on purpose.

Also, when things are going well, many mums find it useful to reflect on their feelings about their baby. If you feel like things aren’t going how you want them to your Health Visitor can help you. We are asking all mums in your area to complete the questionnaire. For now, we are only asking biological mums who are the main carers of their new baby.

#### Why are you asking these questions?

In partnership with the Department of Health Sciences at the University of York, we are exploring how useful these questions are in showing when relationships between mum and baby are going well and not so well. You don’t have to answer these questions if you don’t want to, and you can stop completing the questionnaire at any time – your decision will not affect your relationship with your Health Visitor or the support they offer you.

#### What will happen to my answers?

Your Health Visitor will look at your answers and talk to you about your relationship with your baby. Health Visitors are highly trained and understand that being a mum is different for everyone.

If the questionnaire is useful, it may help Health Visitors in offering future support and training to parents around forming a good relationship with their baby. Your answers will be shared with colleagues in the Department of Health Sciences at The University of York (the university are partnering with Rotherham Doncaster and South Humber NHS Foundation Trust to explore the usefulness of the questionnaire).

##### 1.1.1 How will we use information about you?

1.1.2 Your NHS Trust will not share any identifiable information about you (e.g. your name or address) with the University of York. The University will examine all anonymous answers on the Me and My Baby questionnaire to see if the questions are helpful in identifying where the relationship between mums and their new baby may be difficult or where some support may be helpful. These findings could help to improve the care new mums across your area receive in the future. The findings will be shared in reports, copies of which will be available on the following websites:

- If you live in xxxxx see xxxxx
- If you live in xxxxx see xxxxx
- Research team website: https://www.arc-yh.nihr.ac.uk/home

The research team at the University of York will only have access to <u>fully</u> anonymised data, they will not receive any data or codes that can be used to identify you and they will not be able to see your name or contact details.

The research team will keep all data safe and secure on University of York servers. Once we have finished the study, the University of York will keep the fully anonymised data for 10 years at which point it will be securely destroyed.

##### 1.1.3 What are your choices about how your information is used?

⍰ 11 You can stop being part of the study at any time, without giving a reason, but we will keep information about you that we already have.
⍰ 11 We need to manage your records in specific ways for the research to be reliable. This means that we won’t be able to let you see or change the data we hold about you.

##### 1.1.4 Where can you find out more about how your information is used?

*You can find out more about how we use your information*

⍰ at www.hra.nhs.uk/information-about-patients/
⍰ by asking a member of the research team
⍰ The sponsor for this study is the University of York https://www.york.ac.uk/staff/research/governance/research-policies/policy-for-clinical-research
⍰ at the University of York data protection officer’s website: https://www.york.ac.uk/records-management/dp/
⍰ by ringing your Health visiting service on the numbers below

**If you would like more information, please contact your Health Visiting service in XXXXX on XXXXX, or XXXXX on XXXXX**

## Appendix 2 The MaMB. NOTE: this measure is under further development. Please contact the corresponding author if you wish to use it in the format below

### Me and My Baby

Having a new baby can bring up lots of different feelings and emotions. This questionnaire is designed to explore how you are feeling about being a parent to your baby.

Answering these questions will help us to understand how things are going for you. There is space on the back of this page for you and your Health Visitor to explore why you have answered the way you have if you wish. **Thinking about your feelings about your baby, choose the response for each statement that feels right to you…**

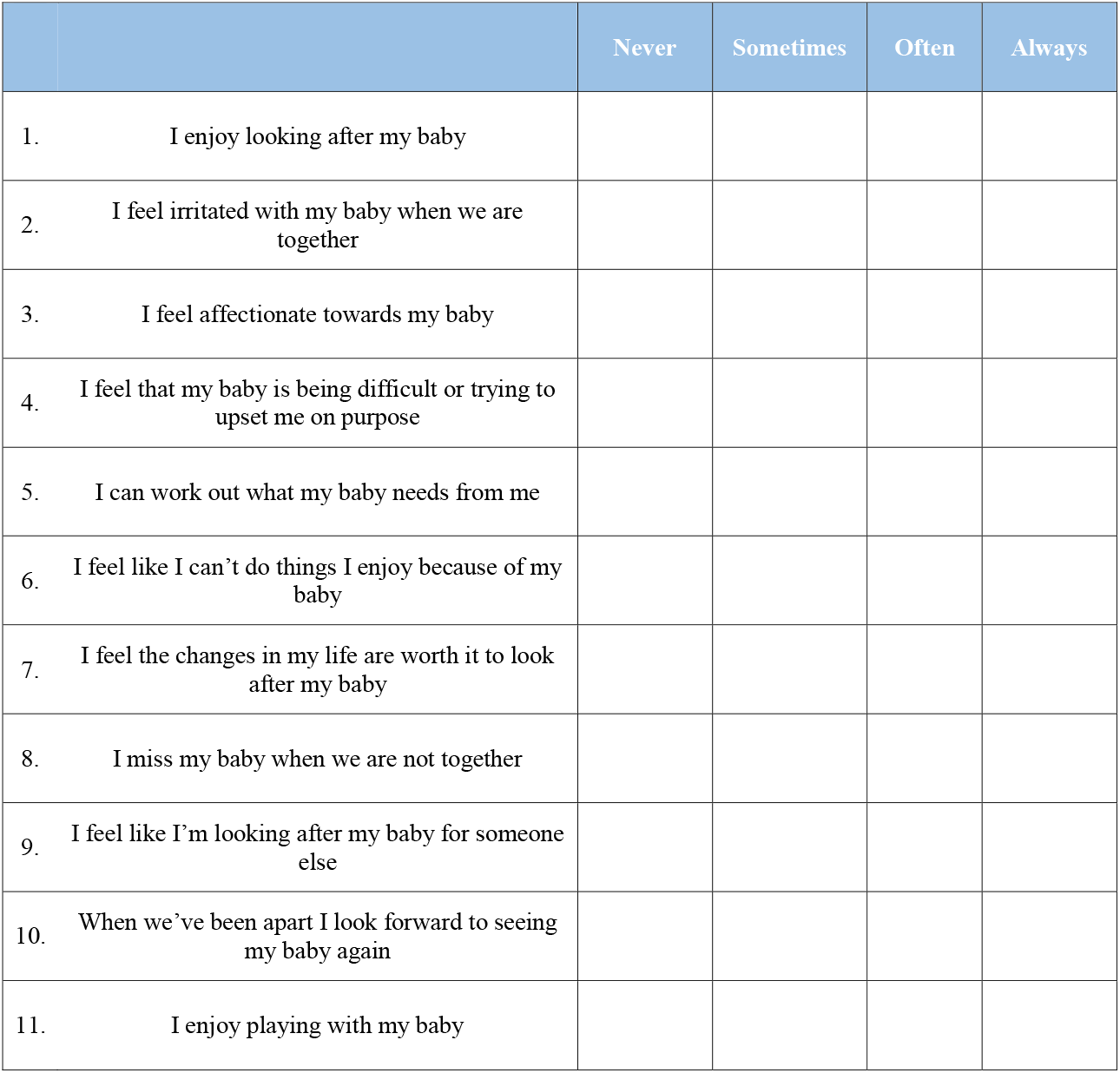
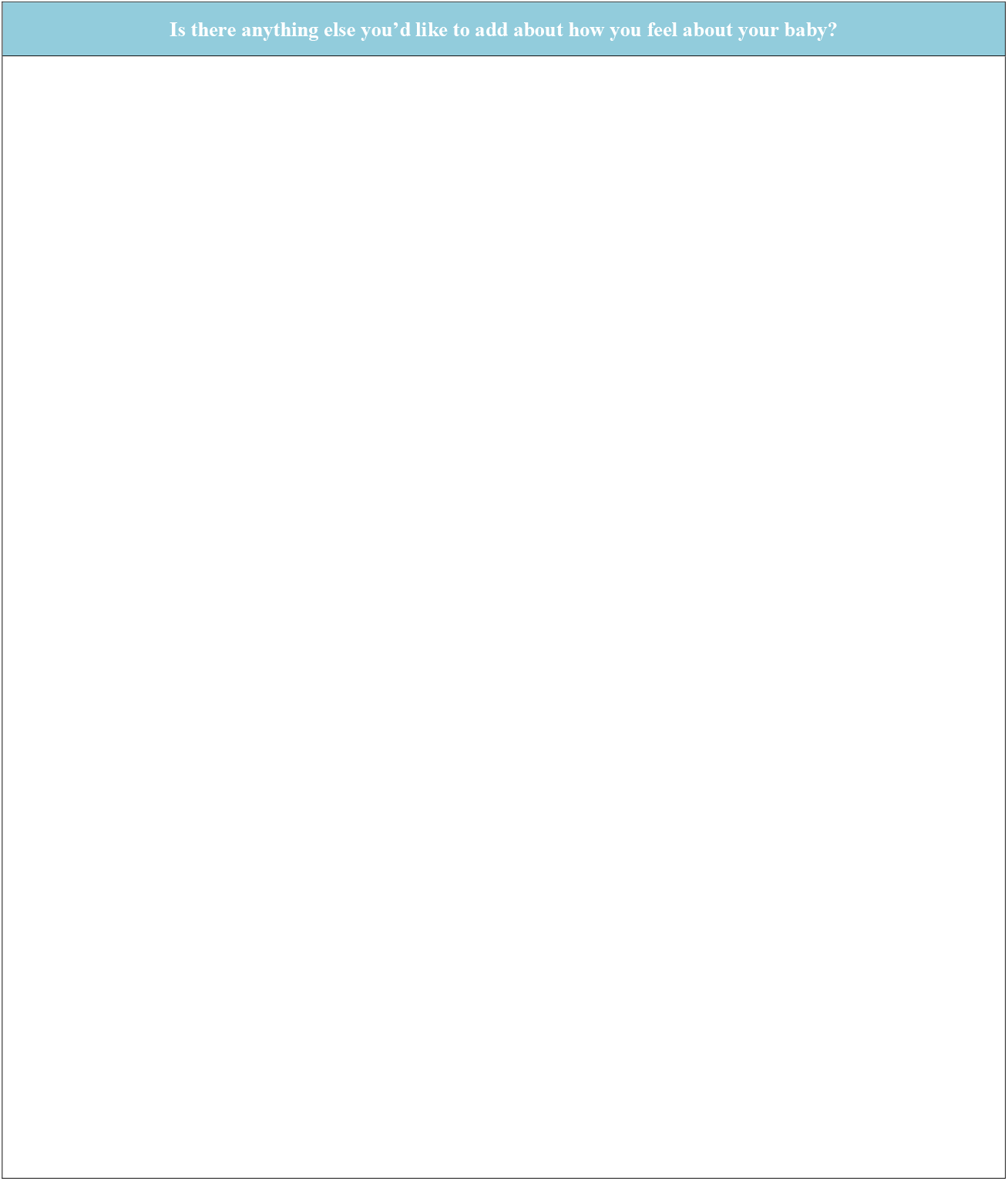

#### Scoring Sheet

The score for each response is in the equivalent box below – find the option selected by the parent for each question and add up the scores. Higher scores on this tool suggest that a parent is finding it difficult to develop an appropriate bond with their infant. **It is important to note that there are no validated cut offs for clinical concern on this tool – so combine scores with your professional judgement when deciding what to do next for a parent**.

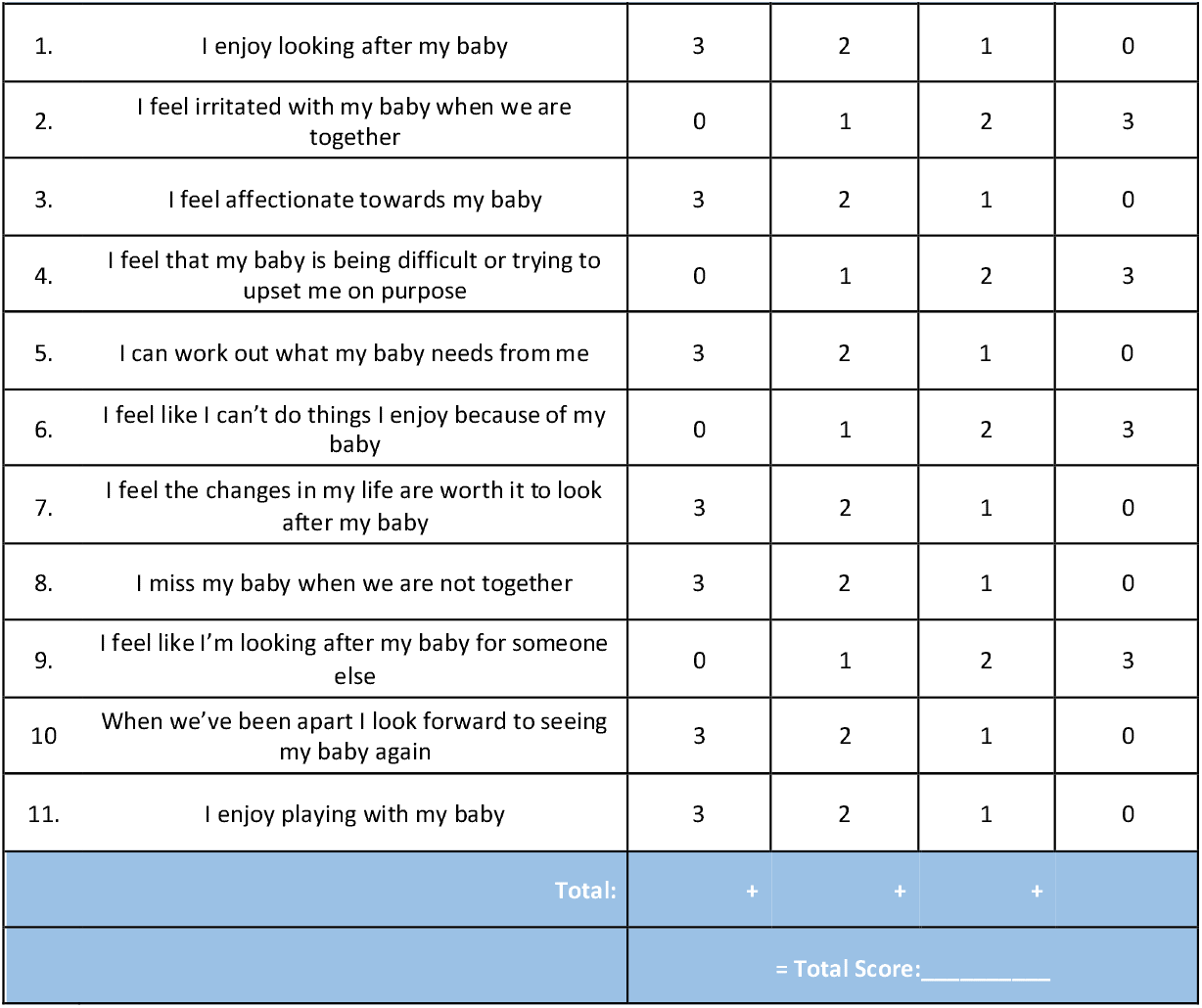

**Figure S1.**
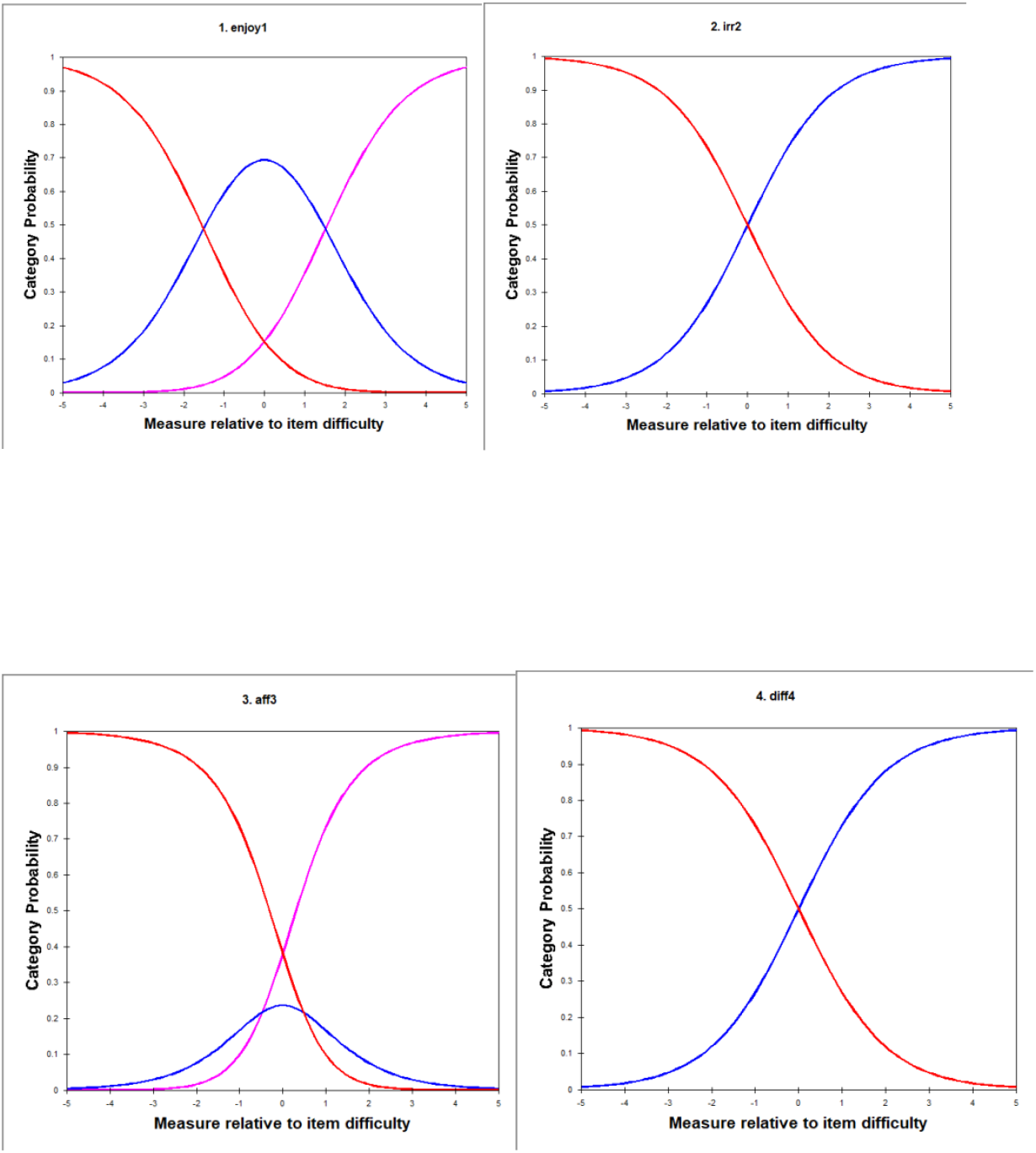
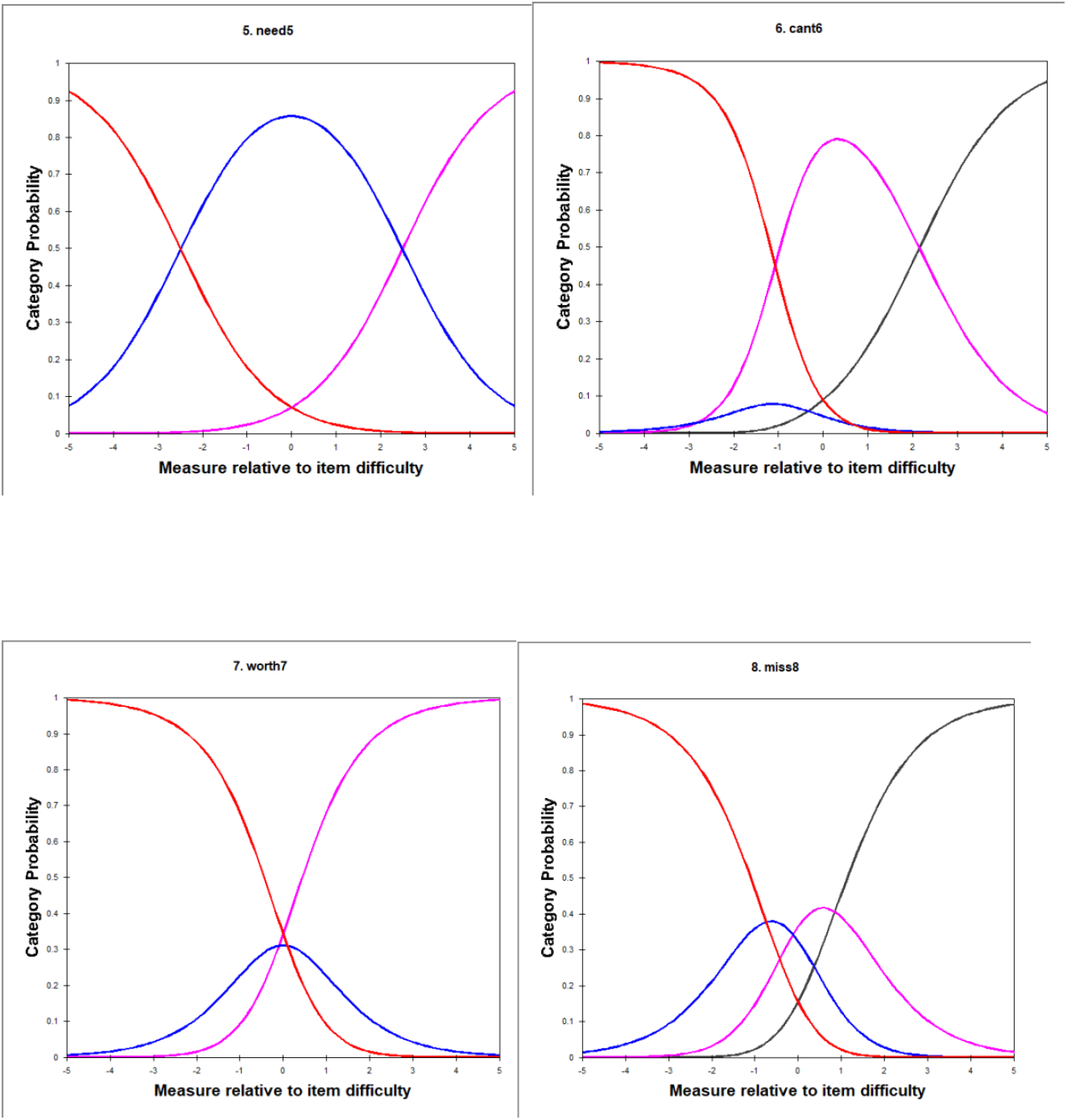
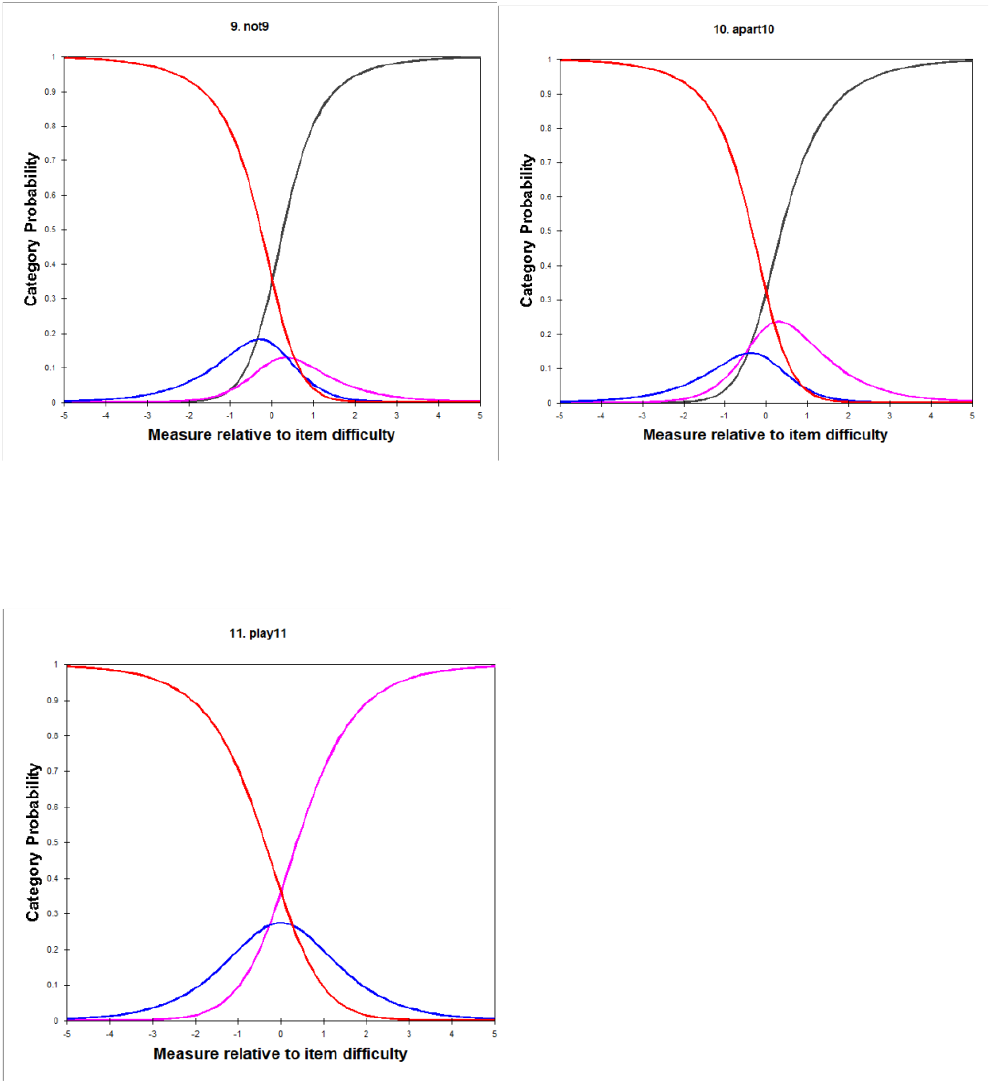
Category probability plots for the 11 items of the MaMB scale.

**Figure S2.**
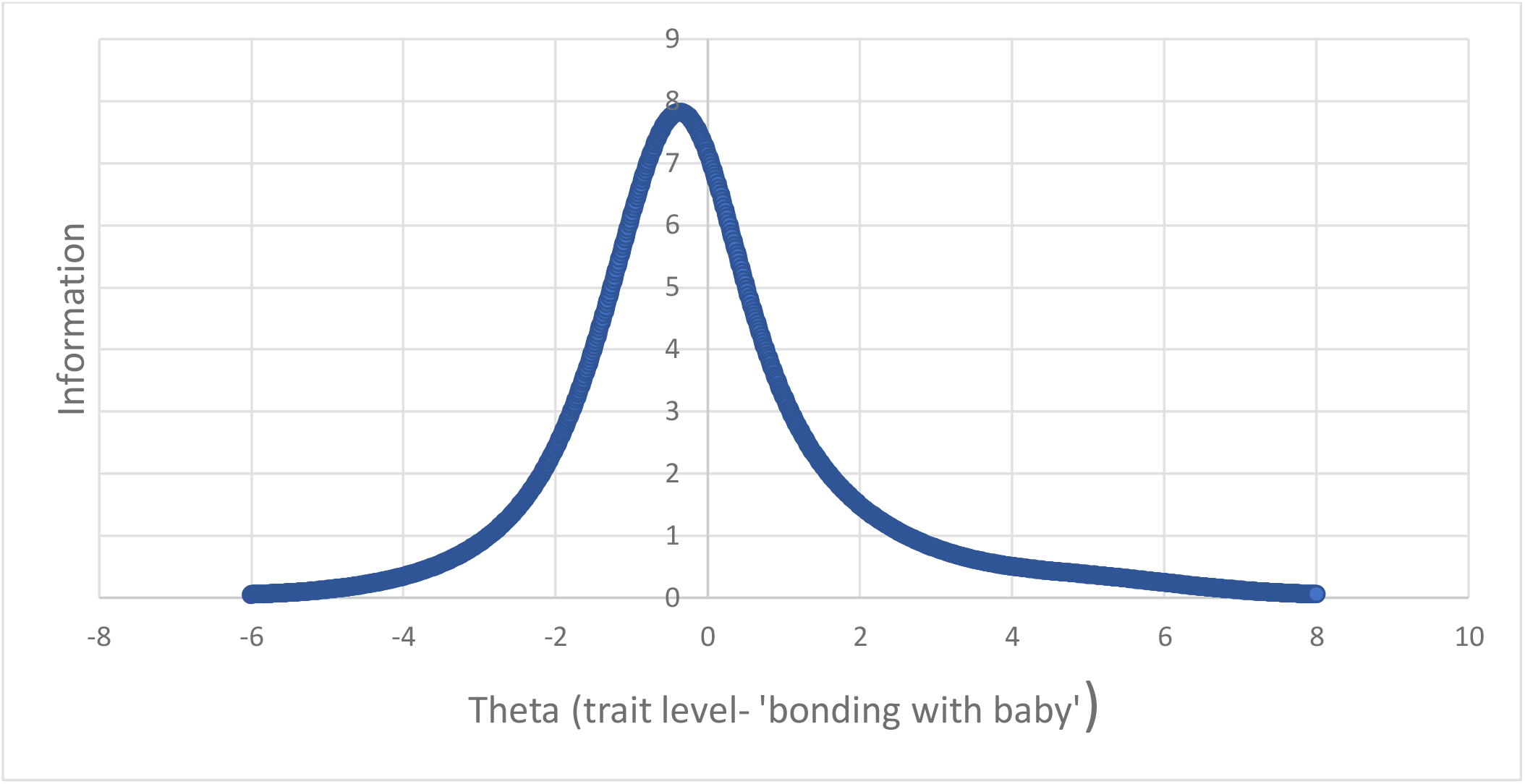
Test Information curve for the MaMB questionnaire

